# Motor network activation and white matter connectivity patterns are related to clinical outcomes following perinatal and childhood arterial ischemic stroke

**DOI:** 10.1101/2024.12.09.24317592

**Authors:** SR Curnow, J Chen, AEL Warren, R Beare, MT Mackay, JYM Yang

**Author notes:** Corresponding author: Sarah Curnow; The Royal Children’s Hospital, 50 Flemington Road, Parkville, Victoria, Australia, 3052.

## Abstract

**Background:** Motor outcomes following perinatal and childhood stroke are highly variable and likely reflect complex structural and functional changes within motor-related brain networks. Greater understanding of these network mechanisms is needed to improve patient-specific prognostication and develop targeted therapies to restore motor function. This study explored post-stroke motor-related reorganization patterns through assessment of functional MRI (fMRI) activation and white matter connectivity patterns.

**Hypothesis:** We hypothesized that activation and connectivity patterns would differ between children with and without hemiparesis following stroke.

**Methods:** Twelve children (age 9-19 years) with a history of perinatal or childhood stroke underwent 3T MRI with diffusion-weighted imaging and task-related hand-motor fMRI. The Paediatric Stroke Outcome Measure was used to assess motor outcome and dichotomized into two groups. MRI data were processed and analyzed for whole-brain fMRI activation patterns and white matter track-weighted functional connectivity (TW-FC) profiles, in relation to motor outcomes.

**Results:** Divergent fMRI activation and TW-FC patterns were seen between the two groups. Children in the no hemiparesis group were more likely to retain an ipsilesional fMRI activation pattern while children in the hemiparesis group were more likely to demonstrate bilateral or contralesional patterns. Increased utilization of cortical-subcortical pathways was identified by TW-FC in both groups. Greater TW-FC strength when moving the stroke-affected hand was also seen in the no hemiparesis group compared to the hemiparesis group.

**Conclusions:** We identified distinct patterns of motor network activation and white matter connectivity relating to clinical outcomes after perinatal and childhood stroke. These patterns have potential to improve prognostic ability and aid identification of infants and children who would most benefit from targeted therapy to improve long-term motor outcomes.

## Introduction

Stroke in childhood is a significant cause of brain injury and can lead to lifelong disability. Long-term neurological outcomes however, are highly variable, with one third of neonates and half of children developing a hemiplegia,^1^ and epilepsy, language difficulties and behavioral problems occurring in both groups.^1^ Understanding the factors that lead to this range of outcomes is key to providing appropriate prognostic counselling and identifying high risk children for targeted early rehabilitation interventions.^2,3^

Advances in MRI-based neuroimaging techniques provide opportunities to study the brain as functionally and structurally connected networks and the impact of stroke on these networks.^4^ Techniques include blood-oxygen-level-dependent (BOLD) functional MRI (fMRI), used to investigate brain functional networks, and diffusion MRI tractography, which interrogates brain white matter (WM) organization. The track-weighted functional connectivity (TW-FC) map^5^ uniquely employs both techniques to combine tractography reconstructed at a whole-brain level and voxel-wise fMRI activation, providing novel insights into both the pattern and strength of the structural connections that underpin the functional network.

Key aspects of motor network recovery studied in perinatal and, to a lesser degree, childhood stroke, include connections between ipsilesional cortical and subcortical structures,^6,7^ inter-hemispheric connections via the body of corpus callosum,^8,9^ the role of the contralesional hemisphere ^10,11^ and the structural integrity of ipsilesional corticospinal tract (CST).^12,13^ No single imaging feature has been shown to be a reliable predictor of post-stroke motor outcomes and limitations conceptualizing stroke recovery mechanisms persist. Moreover, while fMRI and diffusion tractography have been utilised for the purpose of investigation post-stroke recovery, TW-FC has not been applied to this setting.

The objective of this study was to investigate patterns of motor network reorganization following perinatal and childhood stroke and how they relate to motor outcomes at chronic stroke recovery using a novel imaging technique. We aimed to compare the patterns of hemispheric laterality of BOLD fMRI hand-motor task activation and of TW-FC maps between children with and without persistent hemiparesis after perinatal and childhood arterial ischemic stroke (AIS).

## Methods

### Participants

Term-born children with radiologically confirmed AIS, diagnosed in the perinatal, infant or childhood period were recruited from three clinical registries (Stroke, Epilepsy and Rehabilitation Programs) at the Royal Children’s Hospital, Australia. Inclusion criteria included the ability to undergo an awake MRI and a stroke involving the middle cerebral artery territory. The study was approved by the hospital ethics committee and written informed parent consent was obtained.

### Clinical assessment

A standard neurological examination was performed to confirm the presence or absence of hemiparesis and to score participants using the Pediatric Stroke Outcome Measure (PSOM).^14^ The sensorimotor PSOM was dichotomised such that a score of 0-0.5 categorized either no hemiparesis or hemiparesis without functional deficit (the no hemiparesis group) and a score of 1-2 categorized a hemiparesis with functional deficit (the hemiparesis group). Handedness was ascertained by determining which hand was consistently used for skilled tasks. The hand controlled by the stroke-affected (ipsilesional) hemisphere was labelled the “stroke hand” and the hand controlled by the unaffected (contralesional) hemisphere was labelled the “non-stroke hand”.

### Diagnostic imaging assessment

Where possible, acute MRI was reviewed and the modified pediatric Alberta Stroke Programme Early CT Score (ASPECTS)^15^ was applied to determine stroke extent and distribution.

### Imaging data acquisition

Imaging was performed on a 3T MRI scanner and included diffusion-weighted imaging (DWI), finger or wrist-motor BOLD-fMRI and a 3D T1-weighted anatomical scan. Mock MRI was performed in younger participants to improve acquisition success. The acquisition protocol was as follows: 1) DWI 2.3 mm isotropic voxels, TR/TE = 3.2/0.11s, matrix: 110 x 110, 60 diffusion directions, and b-value = 2800 or 3000 mm/s^2^, and 4 to 7 b = 0 mm/s^2^ volumes; 2) fMRI: 2.5 isotropic voxels or 1.6 x 1.6 x 3 mm voxels, TR/TE = 1.5/0.03s or 3/0.04s, volumes 130 or 65, matrix size = 128 x 128 or 104 x 104); 3) T1: 0.8 or 0.9 mm isotropic voxels, TR/TE = 1.9/0.002s. The motor fMRI paradigm involved an index finger-tapping task using an on-off block design alternating between 15 second active and rest blocks (6 active, 7 rest), totaling 195 seconds. Wrist flexion/extension movement was used if finger movement was limited due to extent of weakness. Patient compliance and any mirror hand movements were monitored and videoed throughout fMRI scanning.

### Imaging processing

The study involved two components: 1) task-based fMRI activation analysis and 2) TW-FC analysis.

Functional MRI data were pre-processed using fMRIPrep software version 20.1.1.^16^ This involved corrections for geometric distortion, slice-timing, and head-motion, alignment of the fMRI data to the T1-weighted scan^17^ and Gaussian spatial smoothing (full-width-at-half-maximum=5 mm).^18^ A non-linear warp was calculated between each participant’s MRI and Montreal Neurological Institute-152 6^th^-generation template space.

Diffusion MRI data was pre-processing using MRtrix3 (version 3.0.2-66-g89476794, www.mrtrix3.org).^19^ This involved corrections for thermal noise,^20^ Gibbs-ringing artefacts,^21^ eddy current and motion-induced distortions,^22,23^ geometric distortions^22^ and bias field induced inhomogeneity.^24^ Tissue-specific response functions for grey matter, white matter (WM) and cerebrospinal fluid were estimated using a single-shell, three-tissue algorithm.^25^ WM fibre orientation distributions (FOD) were estimated and reconstructed using multi-tissue Constrained Spherical Deconvolution (MSMT-CSD).

### Study component 1: Task-based fMRI processing

For each participant, BOLD signal changes during the hand-motor task were examined using a general linear model (GLM)-based approach, implemented within fMRI Expert Analysis Tool (FEAT) software version 6.0.4.^26^ This analysis was performed in each participant’s native T1-weighted MRI space, for the left- and right-hand tasks separately. BOLD signal changes were modeled using a boxcar function representing the active and rest task blocks, convolved with the double-gamma hemodynamic response function provided in FEAT, as well as its first-order temporal derivative.^26^ As covariates, 6 head motion parameters were included in the GLM. For statistical inference, contrasts were performed to compare the task and rest conditions. The resulting Z-statistic images were assessed using a voxel-wise uncorrected threshold of Z>3.1 (equivalent to p<0.001).

### Study component 2: TW-FC processing

To create a TW-FC map, whole brain tractography was reconstructed from each participant’s DWI data, using a probabilistic algorithm^27^ and co-registered with their fMRI activation map (Figure 1A). Twenty million streamlines were retained using an FOD cutoff of 0.07 and each streamline was weighted by the average level of fMRI activation in brain areas the streamline traversed. All other steps were as published.^28^ The resultant TW-FC maps were then thresholded at the median intensity value to remove regions containing less probable (i.e. bottom 50%) FC activations.

**Fig 1.**
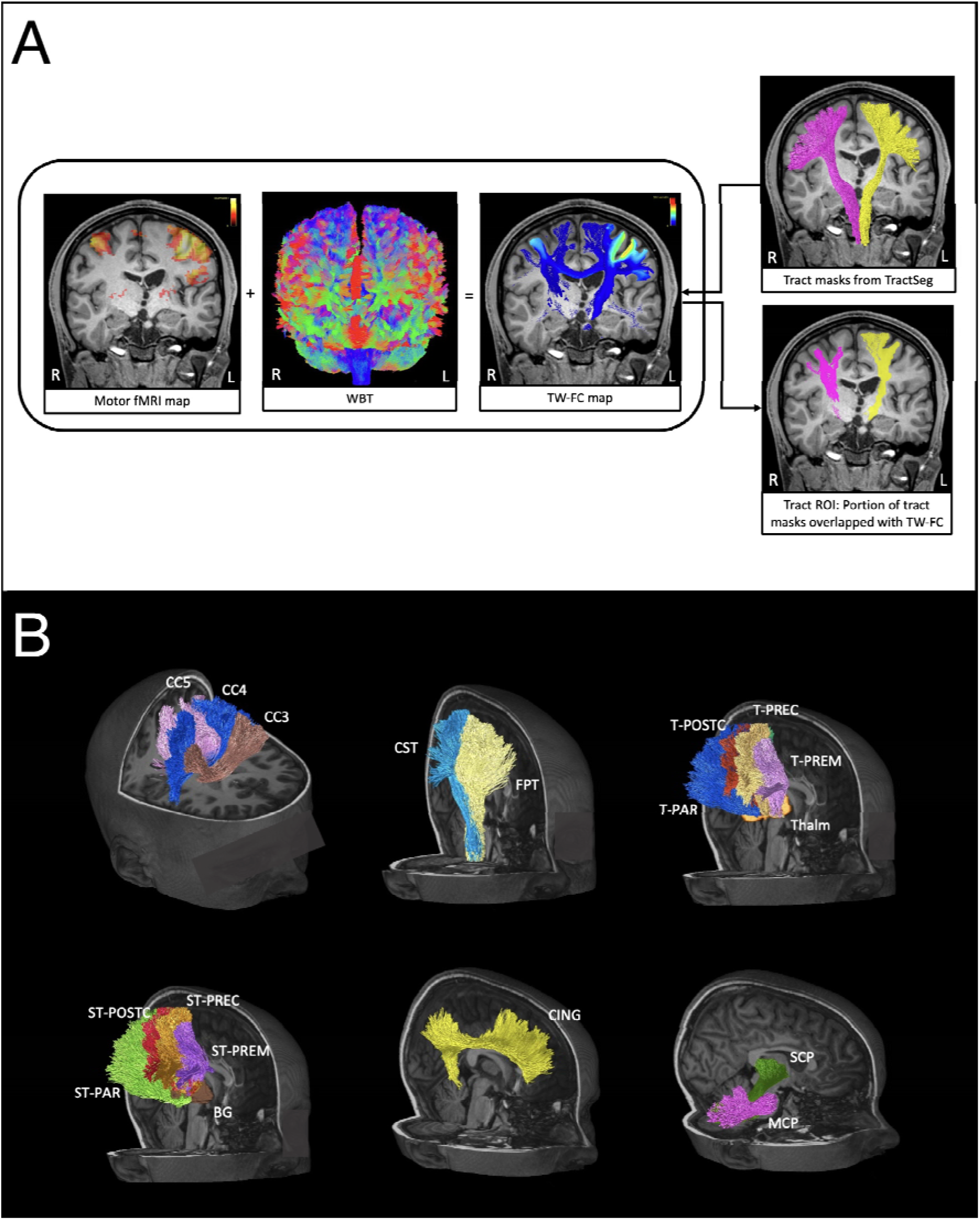
**Panel A: TW-FC data processing pipeline**. *fMRI: functional MRI; L: left; R: right; ROI: regions of interest; TW-FC: Track weighted-functional connectivity; WBT: whole brain tractogram.* **Panel B: Pre-selected sensorimotor network white matter tracts**. This includes the corpus callosum (A), projection tracts (corticospinal tract, CST and fronto- pontine tract, FPT; B), thalamocortical tracts (C), striatocortical tracts (D), Cingulum (CING; D), and cerebellar motor and sensory tracts contained in superior cerebellar peduncle, SCP and middle cerebellar peduncle, MCP, respectively (F). BG: basal ganglia; L/R: Left/Right; MCP: middle cerebellar peduncle; ST_PAR: striato-parietal; ST_PREC: striato-precentral; ST_PREM: striato-premotor; ST_POSTC: striato-postcentral; STR: superior thalamic radiation; T_PAR: thalamo-parietal; T_PREC: thalamo-precentral; T_PREM: thalamo-premotor; T_POSTC: thalamo-postcentral; Thalm: thalamus

An automated deep-learning based WM tract segmentation tool (TractSeg) was applied to reconstruct 72 major WM tracts from each participant’s DWI data.^29^ Twenty-eight WM tracts known to be implicated in the sensorimotor network were pre-selected (Figure 1A). Binarized masks of the tracts of interest were overlaid on the TW-FC maps to guide classification of WM tracts involved in the TW-FC maps. The sum TW-FC intensity values of all voxels contained within the tract mask, defined as portions of the tract masks overlapping the thresholded TW-FC maps, were used for the calculation of laterality indices (LI) (Figure 1A).

A region-of-interest (ROI) based, quantitative TW-FC laterality analysis was carried out to investigate patterns of TW-FC lateralization. LIs based on the TW-FC values derived from all WM tract ROI pairs were calculated using the following equation:

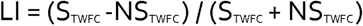

STWFC is the sum TW-FC value derived from stroke (ipsilesional) tract ROI; and NSTWFC is the corresponding value derived from the non-stroke (contralesional) tract ROI. The LI ranges between -1 to 1. Positive LIs indicate lateralization to the stroke/ipsilesional hemisphere, negative LIs indicate lateralization to the non-stroke/contralesional hemisphere, and LIs close to and including zero indicate bi-laterality.

Box plots were used to demonstrate patterns of TW-FC laterality from all participants. The within-participant TW-FC laterality patterns were visually compared between the stroke and the non-stroke hand-motor fMRI tasks. The between-participant TW-FC laterality patterns were visually compared between the no hemiparesis and the hemiparesis groups.

### Data analysis

LIs between participants in both groups were analyzed using a multilevel regression model incorporating participant ID as a random effect. The quantitative LI analysis was supplemented by detailed case-based exploratory assessment of trends using visual analysis. We reviewed all fMRI activation and TW-FC maps, including scrutinizing the extent and pattern of fMRI BOLD activation clusters individually for anatomical localization and the involved WM tracts, as guided by TractSeg outputs.

The corresponding author has full access to all the data in the study and takes responsibility for its integrity and analysis.

## Results

### Participant characteristics

Participants included ten males and two females, ranging from nine- to 19-years-old at time of MRI and clinical assessment (Table 1). Stroke hemisphere was left in eight and right in four. Stroke history was perinatal arterial ischemic stroke (PAIS) in five, presumed perinatal infarction in two and childhood AIS in five. Six had a PSOM 0-0.5 and were placed in the no hemiparesis group and six had a PSOM 1-2 and were placed in the hemiparesis group. Handedness correlated with the unaffected hemisphere in all but two: participant ID2 was right-handed despite a left hemisphere stroke and participant ID4 was ambidextrous.

**Table 1:**
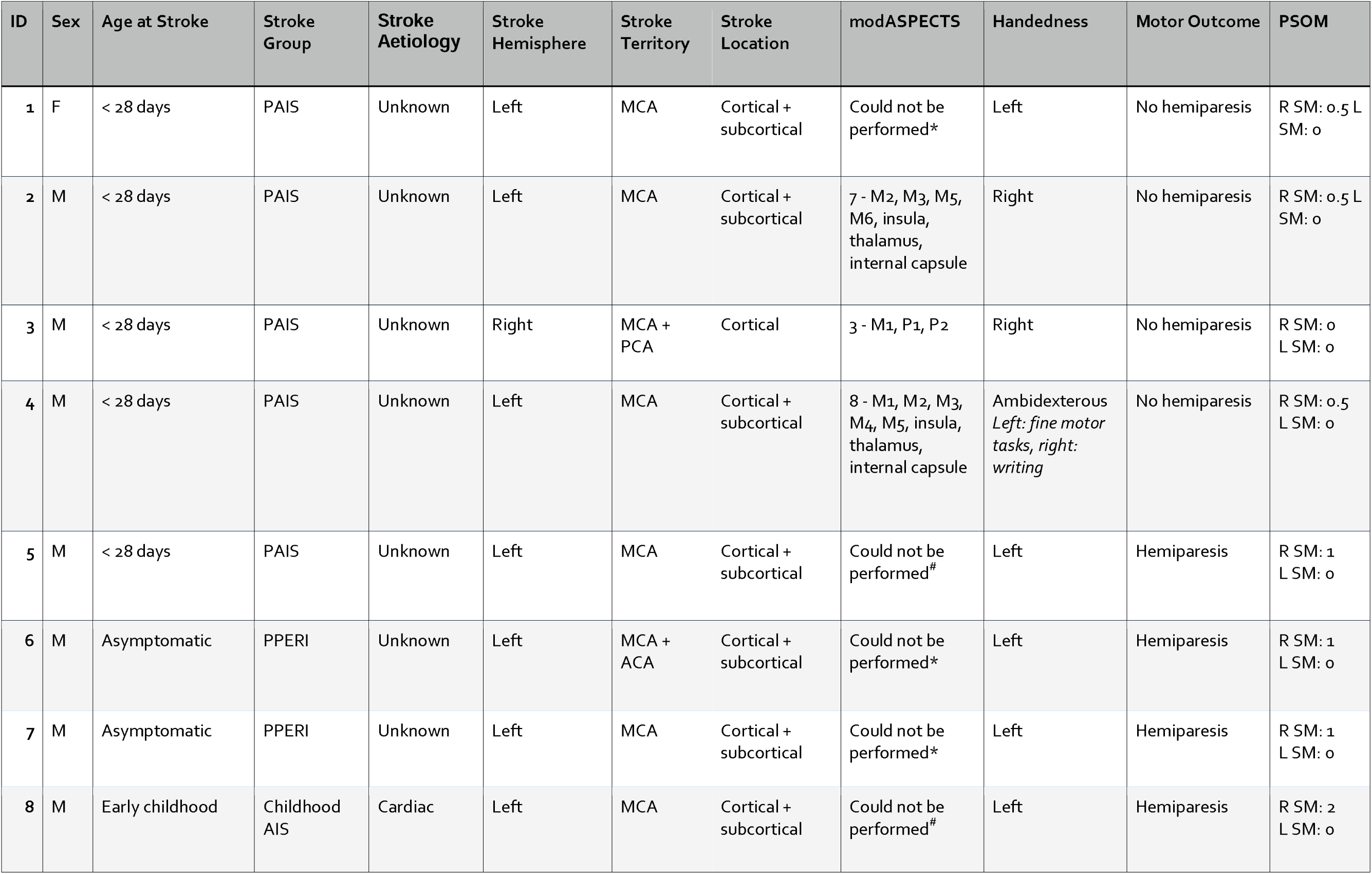
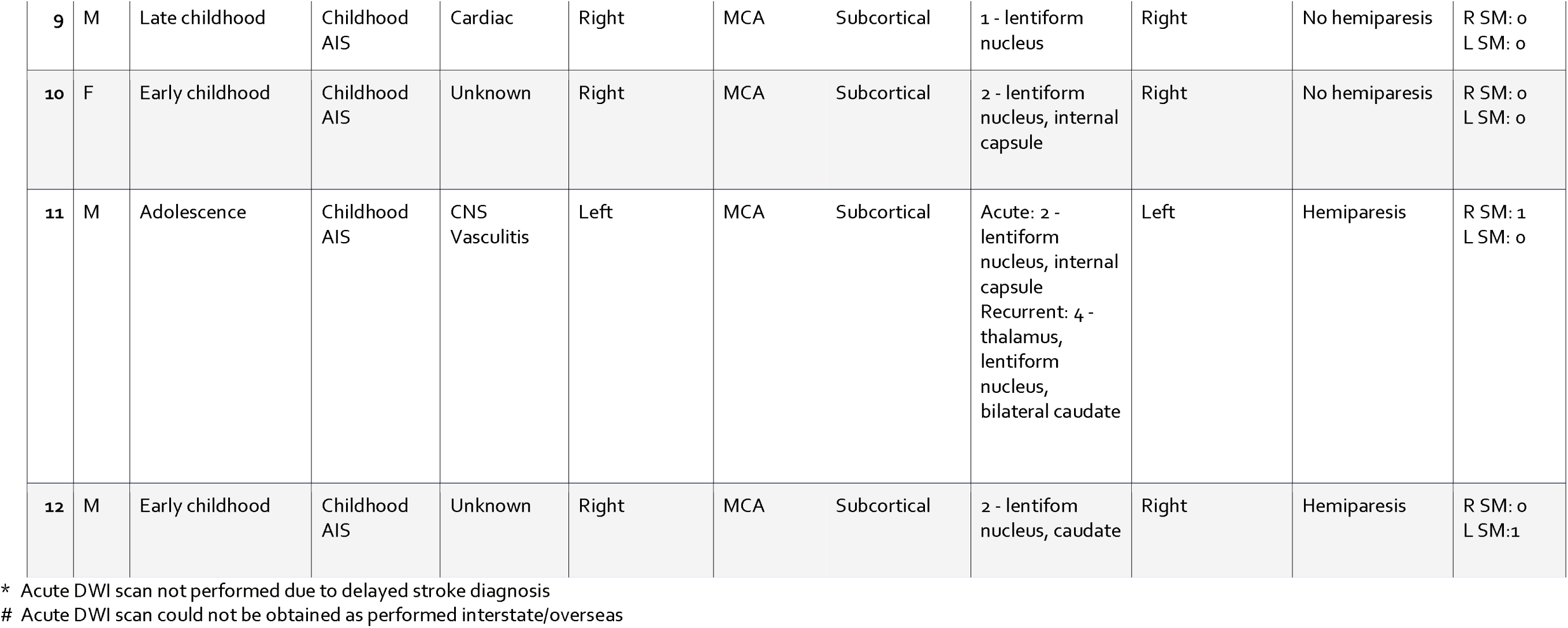
Participant characteristics. ACA: anterior cerebral artery; AIS: arterial ischemic stroke; CNS: central nervous system; d: days; DWI: diffusion weighted imaging; F: female; ID: identification number; L: left; M: male; modASPECTS: modified pediatric Alberta Stroke Program Early CT Score; m: months; MCA: middle cerebral artery; M1-6: MCA territories; PAIS: perinatal arterial ischemic stroke; PCA: posterior cerebral artery; P1-2: PCA territories; PPERI: presumed perinatal infarction; PSOM: Paediatric Stroke Outcome Measure; R: right; SM: sensorimotor; y: years.

| ID | Sex | Age at Stroke | Stroke Group | Stroke Aetiology | Stroke Hemisphere | Stroke Territory | Stroke Location | modASPECTS | Handedness | Motor Outcome | PSOM |
| --- | --- | --- | --- | --- | --- | --- | --- | --- | --- | --- | --- |
| 1 | F | < 28 days | PAIS | Unknown | Left | MCA | Cortical + subcortical | Could not be performed* | Left | No hemiparesis | R SM: 0.5<br>L SM: 0 |
| 2 | M | < 28 days | PAIS | Unknown | Left | MCA | Cortical + subcortical | 7 - M2, M3, M5, M6, insula, thalamus, internal capsule | Right | No hemiparesis | R SM: 0.5<br>L SM: 0 |
| 3 | M | < 28 days | PAIS | Unknown | Right | MCA + PCA | Cortical | 3 - M1, P1, P2 | Right | No hemiparesis | R SM: 0<br>L SM: 0 |
| 4 | M | < 28 days | PAIS | Unknown | Left | MCA | Cortical + subcortical | 8 - M1, M2, M3, M4, M5, insula, thalamus, internal capsule | Ambidexterous<br>*Left: fine motor tasks, right: writing* | No hemiparesis | R SM: 0.5<br>L SM: 0 |
| 5 | M | < 28 days | PAIS | Unknown | Left | MCA | Cortical + subcortical | Could not be performed<sup>#</sup> | Left | Hemiparesis | R SM: 1<br>L SM: 0 |
| 6 | M | Asymptomatic | PPERI | Unknown | Left | MCA + ACA | Cortical + subcortical | Could not be performed* | Left | Hemiparesis | R SM: 1<br>L SM: 0 |
| 7 | M | Asymptomatic | PPERI | Unknown | Left | MCA | Cortical + subcortical | Could not be performed* | Left | Hemiparesis | R SM: 1<br>L SM: 0 |
| 8 | M | Early childhood | Childhood AIS | Cardiac | Left | MCA | Cortical + subcortical | Could not be performed<sup>#</sup> | Left | Hemiparesis | R SM: 2<br>L SM: 0 |
| 9 | M | Late childhood | Childhood AIS | Cardiac | Right | MCA | Subcortical | 1 - lentiform nucleus | Right | No hemiparesis | R SM: 0<br>L SM: 0 |
| 10 | F | Early childhood | Childhood AIS | Unknown | Right | MCA | Subcortical | 2 - lentiform nucleus, internal capsule | Right | No hemiparesis | R SM: 0<br>L SM: 0 |
| 11 | M | Adolescence | Childhood AIS | CNS Vasculitis | Left | MCA | Subcortical | Acute: 2 - lentiform nucleus, internal capsule<br>Recurrent: 4 - thalamus, lentiform nucleus, bilateral caudate | Left | Hemiparesis | R SM: 1<br>L SM: 0 |
| 12 | M | Early childhood | Childhood AIS | Unknown | Right | MCA | Subcortical | 2 - lentiform nucleus, caudate | Right | Hemiparesis | R SM: 0<br>L SM: 1 |

\* Acute DWI scan not performed due to delayed stroke diagnosis

### Study component 1: Task-based fMRI activation analysis

Divergent BOLD fMRI activation patterns were evident between the no hemiparesis and hemiparesis groups. Children in the no hemiparesis group demonstrated typical contralateral/ipsilesional activation patterns when moving the stroke-affected hand. Children in the hemiparesis group however, demonstrated atypical patterns with either bilateral or contralesional activation on moving the stroke hand. The pertinent findings are illustrated by three participants’ fMRI activation maps (Figure 2).

**Figure 2:**
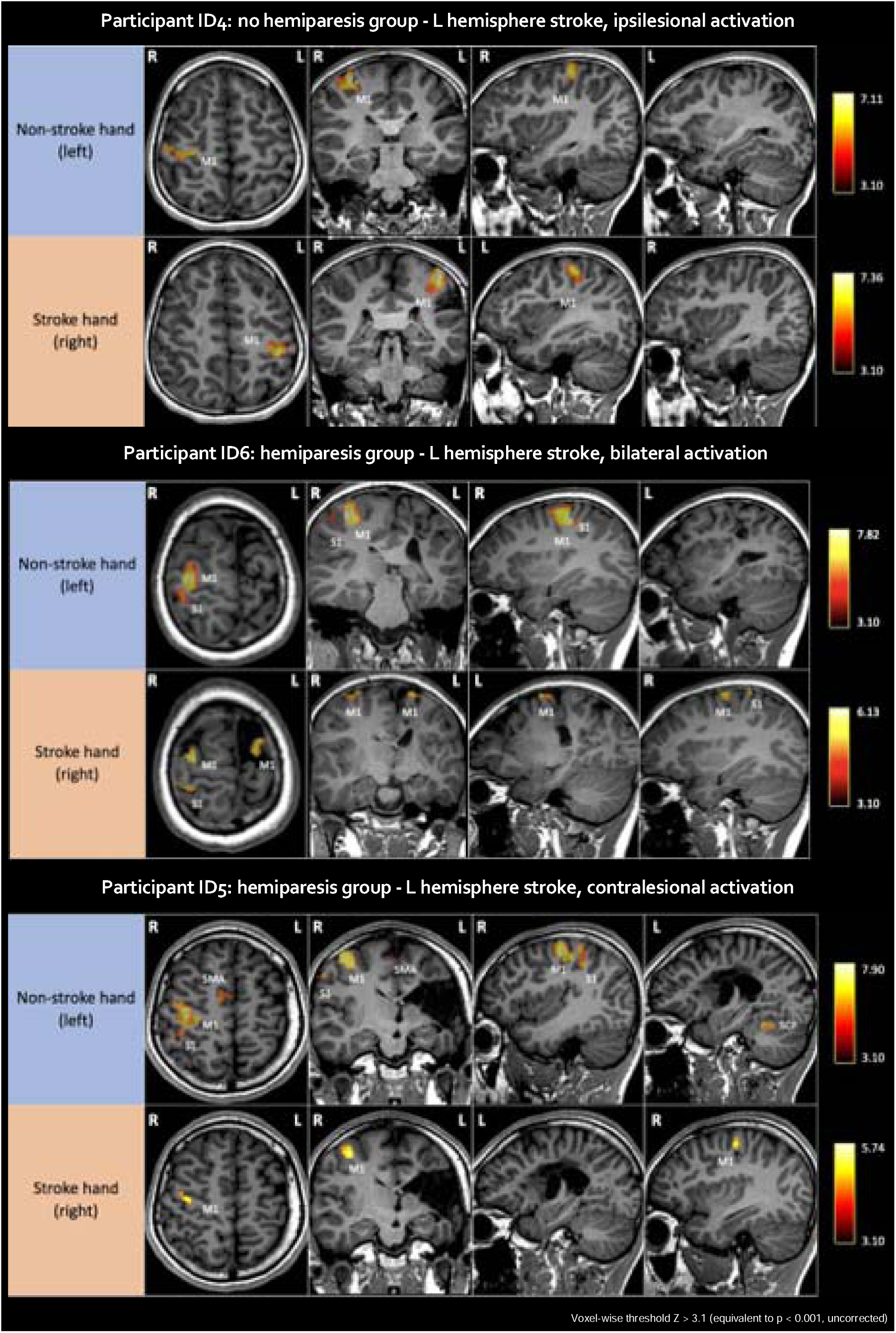
Labelled fMRI activation maps for selected participants. L: left; M1: primary motor area; R: right; S1: primary somatosensory area; SCP: superior cerebellar peduncle; SMA: supplementary motor area. 1st column: axial view; 2nd column: coronal view; 3rd & 4th columns: sagittal view (hemisphere marked). Images displayed using radiological convention. Top two rows: participant ID4 (no hemiparesis group) showing a typical, ipsilesional activation pattern when moving the stroke hand; Middle two rows: participant ID6 (hemiparesis group) showing a bilateral activation pattern when moving the stroke hand; Bottom two rows: participant ID5 (hemiparesis group) showing transfer of function to the contralesional hemisphere when moving the stroke hand.

### Study component 2: TW-FC analysis

Figure 3 shows the TW-FC maps with labelled WM tracts for stroke and non-stroke hand movement for the same three participants. The patterns mirror those seen on the fMRI activation maps. Increased utilization of non-CST pathways, such as the striatocortical and thalamocortical tracts, was also observed in response to movement of the stroke hand in both groups, best demonstrated by participant ID4’s TW-FC map. Furthermore, in the no hemiparesis group, the TW-FC strength was greater when moving the stroke versus the non-stroke hand. A reversed pattern was observed in the hemiparesis group, with greater TW-FC strength seen when moving the non-stroke versus the stroke hand.

**Figure 3:**
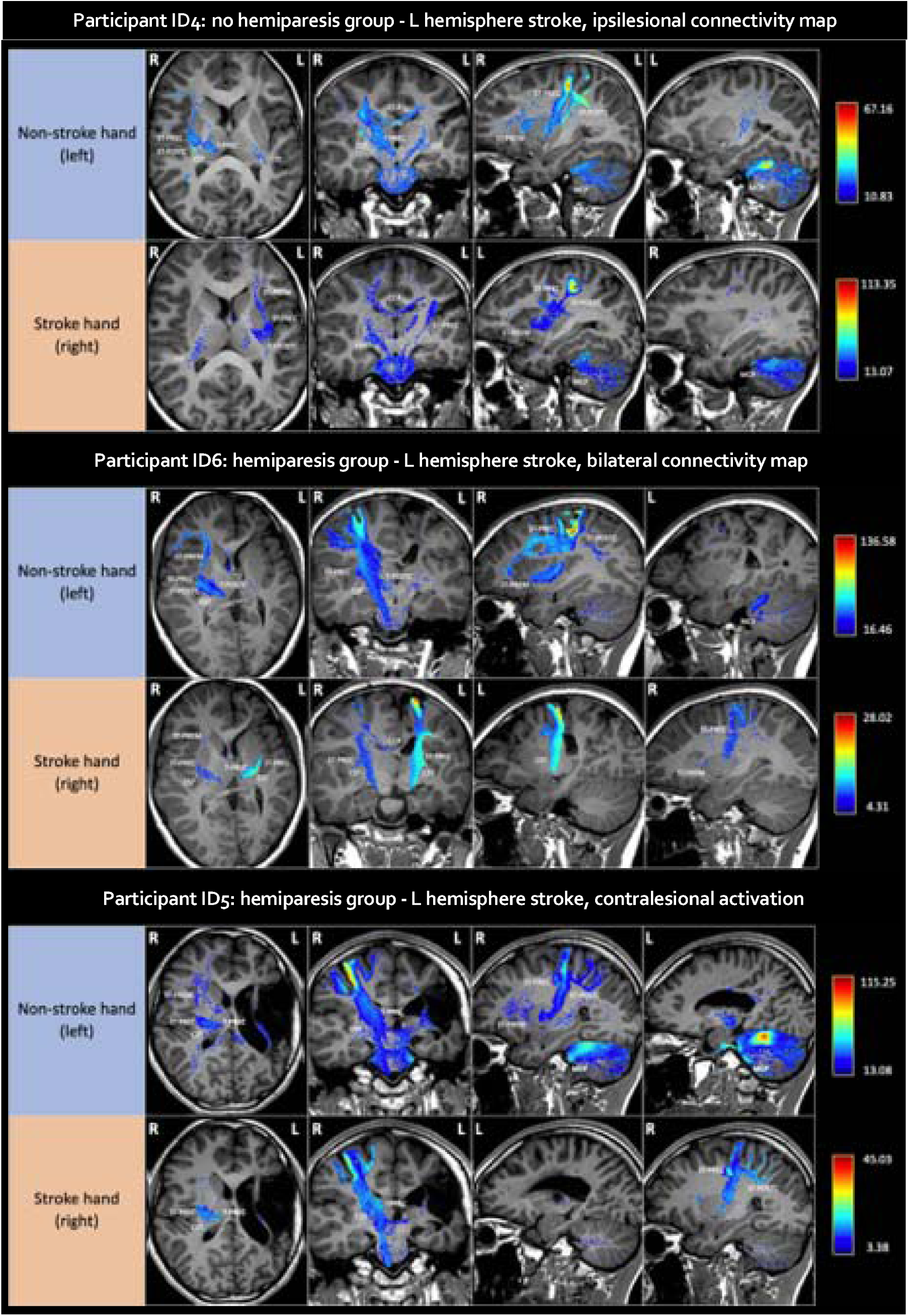
Labelled TW-FC maps for selected participants. CC4: corpus callosum 4 (interhemispheric connectivity between homologous M1 and S1); CST: corticospinal tract; ID: identification number; L: left; MCP: middle cerebellar peduncle; R: right; ST_PREC: striato-precentral; ST_PREM: striato-premotor; ST_POSTC: striato-postcentral; T_PREC: thalamo-precentral; T_POSTC: thalamo-postcentral. 1st column: axial view; 2nd column: coronal view; 3rd & 4th columns: sagittal view (hemisphere marked). Images displayed using radiological convention. Top two rows: participant ID4 (no hemiparesis group) showing a typical, ipsilesional, connectivity map with increased utilisation of non-CST WM tracts when moving the stroke hand; Middle two rows: participant ID6 (hemiparesis group) showing a bilateral connectivity map when moving the stroke hand; Bottom two rows: participant ID5 (hemiparesis group) showing a contralesional connectivity map when moving the stroke hand.

The fMRI activation and TW-FC maps for the remaining participants are included in the supplementary material.

Figure 4 shows the TW-FC strength LI box-plots for all participants by subgroup. No statistically significant group differences were identified (p = 0.754). The general trend however, was that the TW-FC strength when moving the stroke hand was lateralized to the ipsilesional hemisphere for the no hemiparesis group. In contrast, for the hemiparesis group, the TW-FC strength demonstrated reduced laterality to the ipsilesional hemisphere when moving the stroke hand, with participants ID5 and ID12 completely lateralizing to the contralesional hemisphere. Notable outliers were participants ID1 and ID8.

**Figure 4:**
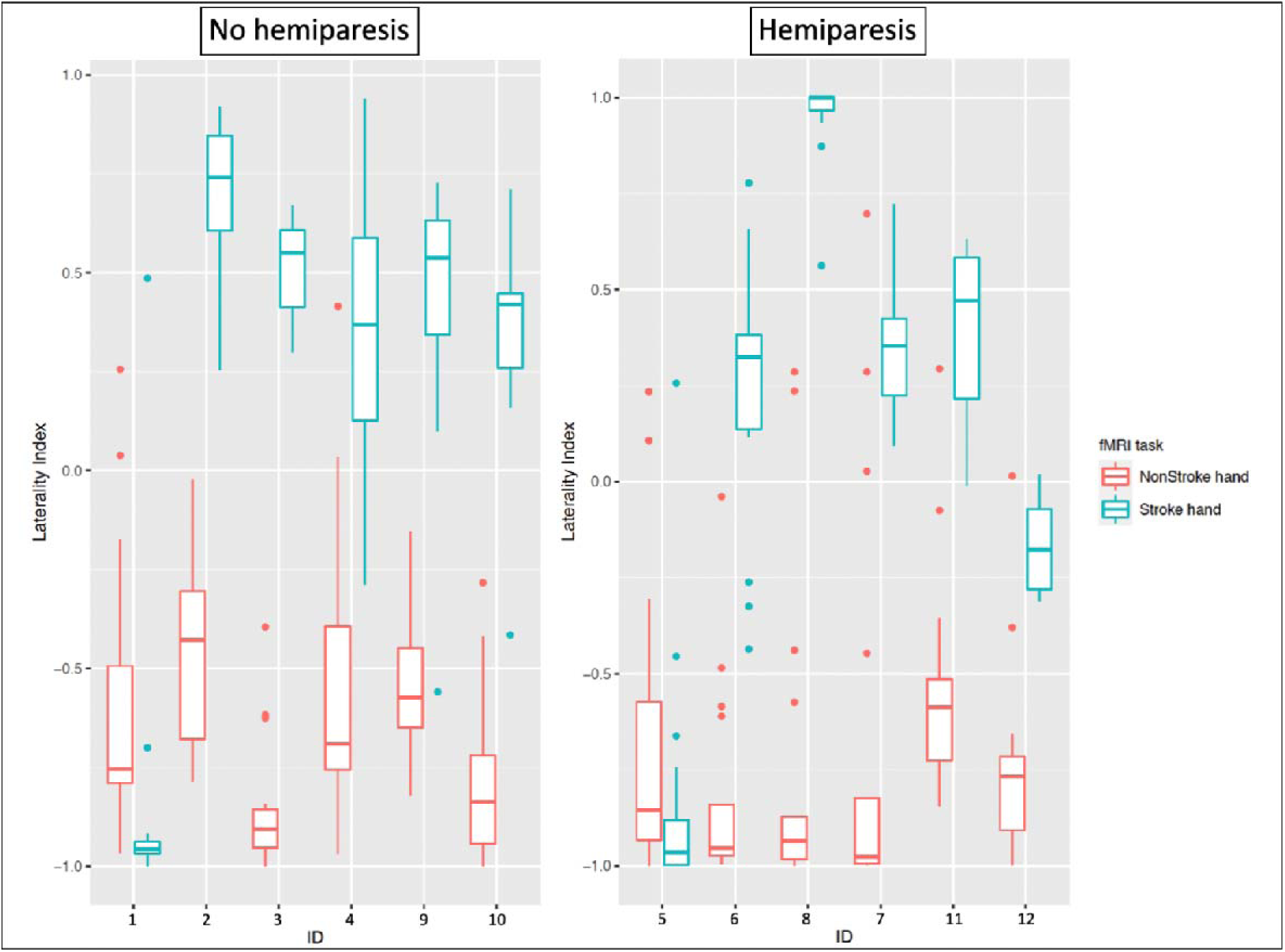
Box-plots showing the laterality index for stroke and non-stroke hand tasks for the no hemiparesis (left) and hemiparesis (right) groups. ID: identification number

## Discussion

We identified different hand-motor fMRI activation and WM connectivity patterns at chronic stroke recovery following perinatal and childhood AIS (Figure 5). Those without hemiparesis/functional impairment had activation and structural connectivity patterns involving functional retention within the stroke-affected hemisphere while those with persistent hemiparesis/functional impairment showed a pattern involving utilization of a greater proportion of the motor network, including the contralesional hemisphere. Regardless of whether or not function was retained within the stroke-affected hemisphere, non-CST motor network WM tracts were employed to move the stroke hand, specifically the striatocortical and thalamocortical tracts. The findings suggest that differences in brain network recovery patterns play a role in the variable motor outcomes seen following perinatal and childhood stroke, with potential clinical implications for prognosis and therapy.

**Figure 5:**
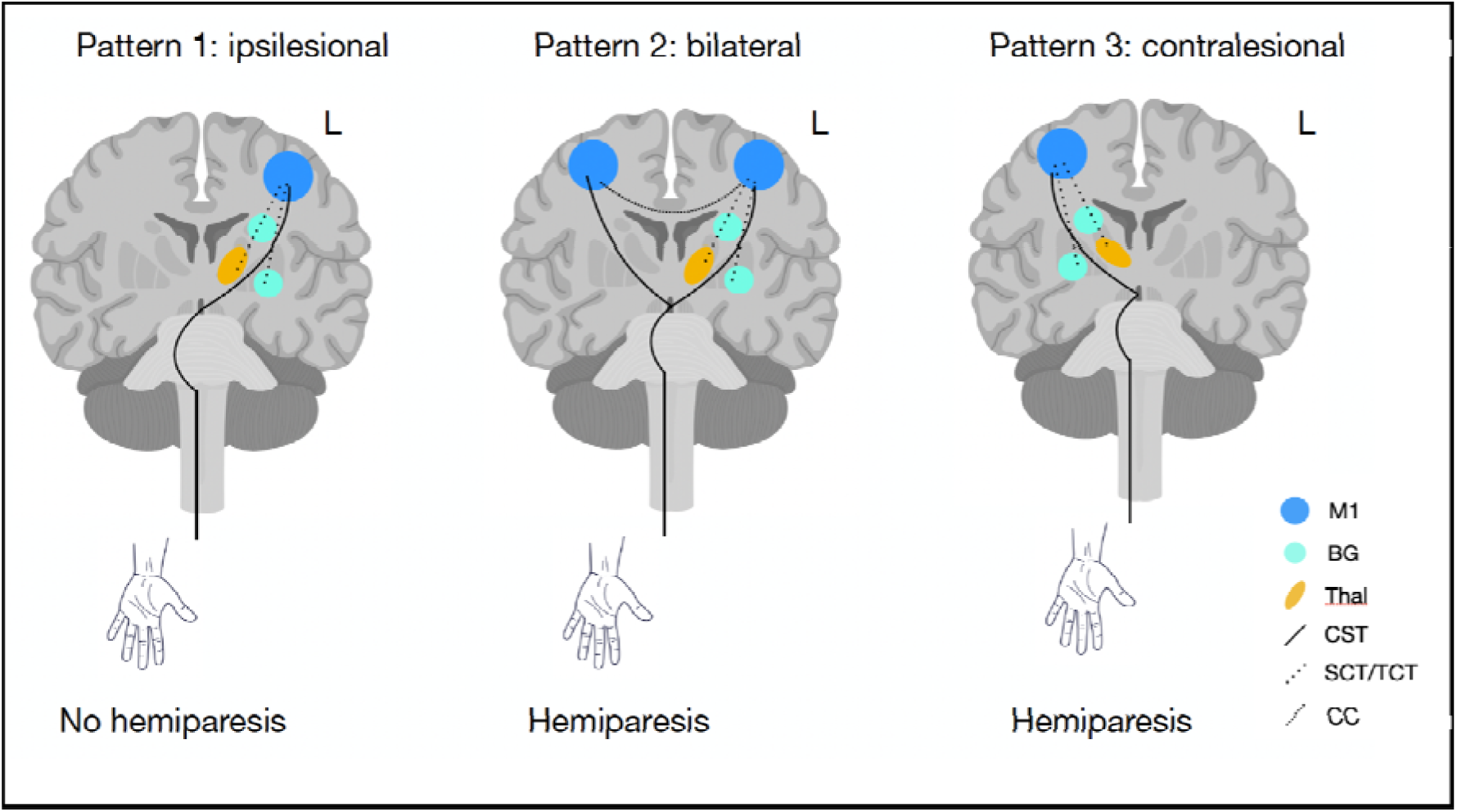
Activation and connectivity patterns on moving the stroke hand following a left hemisphere stroke. L: left; M1: primary motor area; BG: basal ganglia; Thal: thalamus; CST: corticospinal tract; SCT: striatrocortical tract; TCT: thalamocortical tract; CC: corpus callosum

Exceptions to these patterns were participants ID1 (PAIS subtype/no hemiparesis group) and ID8 (Childhood AIS subtype/hemiparesis group). Despite having no functional deficit, participant ID1 demonstrated apparent laterality of stroke hand movement to the contralesional hemisphere. On review of the raw imaging data, the movement obtained during acquisition was below the chosen statistical threshold applied to the fMRI activation maps. When this threshold was altered, a typical ipsilesional activation pattern was indeed demonstrated. In contrast, despite having a persistent hemiparesis with functional deficit, participant ID8’s LI was strongly lateralized to the ipsilesional hemisphere. This participant underwent intensive constraint-based therapy from the time of stroke at age 15 months old. We hypothesise that this may explain the retained motor function within the stroke-affected hemisphere.^30^

Imaging predictors of post-stroke motor outcome have been well-studied in the perinatal setting but less so in the childhood subgroup. In both groups, no single imaging marker has been predictive of long-term outcome. Better motor recovery via retention of function within the stroke-affected hemisphere has been demonstrated in perinatal stroke.^10,31^ The mechanism for this is unclear but may be partially explained by the hypothesis that larger stroke size or injury to key motor network structures may necessitate transfer of function to the unaffected hemisphere. Yet, these atypical patterns are seen following strokes of variable size and location, including both PAIS and periventricular venous infarct imaging phenotypes.^10^ This is also reflected in our study with heterogenous lesion characteristics in both subgroups.

The association between injury to the CST/pre-Wallerian degeneration with poor motor outcomes following both perinatal and childhood stroke has been well-studied.^12,32^ The perinatal subgroup is of particular interest due to bilateral CST projections in the neonatal period.^33^ In children and adults, the majority of CST fibers cross at the level of the pyramids into the lateral aspect of the contralateral spinal cord. A small portion of the fibers (∼ 10%) do not decussate but instead continue into the anterior ipsilateral spinal cord, supplying proximal appendicular and axial muscles.^34^ In the developing fetus, bilateral crossed and uncrossed CST projections are formed, which are then typically withdrawn in a competitive fashion in the first 15-18 months of life,^33^ leading to the typical pattern of contralateral control of function. This process lends itself to the potential for adaptive re-wiring following early brain injury^34^ and the development of compensatory CST trajectories is a key area of interest in the perinatal stroke recovery literature.

Using diffusion tractography, asymmetries in CST projections can be seen as early as 3 months following perinatal stroke^13^ and preservation of the normally-withdrawn ipsilateral CST has been demonstrated at chronic stroke recovery.^10,35,36^ The mechanism is thought to relate to the lack of descending inputs from the lesioned hemisphere reducing the ability of the contralateral CST projections to compete for spinal cord synaptic connections and obtaining their usual dominance.^37^ Degree of involvement of the ipsilesional CST on diagnostic DWI^32,38^ and atrophy of the ipsilesional cerebral peduncles on acute structural imaging^39^ are both indicative of poorer motor outcomes. The alterations in the normal CST developmental process therefore occur early, before a clinical motor deficit is evident, and may represent a therapeutic target for early post-natal intervention via techniques such as transcranial magnetic stimulation.^37,40^

The demonstration of additional non-CST WM tract utilization following perinatal and childhood stroke, is novel to this study. While the important role cortical-subcortical structural relationships play in post-stroke motor network reorganization has been shown, to our knowledge, no previous studies have illustrated such a relationship by employing the TW-FC or other such neuroimaging techniques. The postulated role for these non-CST WM tracts is compensation for the disrupted primary motor/sensory pathways and/or for injured subcortical areas.^41^ For example, topographic displacement of motor function representation from the M1 to the supplementary motor area as demonstrated by task-based fMRI following perinatal stroke^10^ perhaps necessitates recruitment of alternate

descending motor pathways. The functional role of the basal ganglia and thalamus in the control of motor preparation and execution^42,43^ and their structural connectivity to both the M1 and other cortical structures such as the premotor cortex,^44^ provide a means for the potential transfer of function in the case of early M1 injury. The unique finding from this study adds to our understanding of the mechanisms of motor network reorganization after perinatal and childhood stroke.?

The application of advanced neuroimaging techniques using contemporary and rigorous pre-processing and analysis methods is a strength of this study. To our knowledge, there have been no studies employing TW-FC in motor network evaluation following perinatal or childhood stroke. The chosen tractography method, combining the use of an MSMT-CSD model and a probabilistic tracking algorithm, improves the accuracy of WM fibre orientation estimates and anatomical accuracy of the whole-brain tractogram reconstructions.^45^ This technique therefore allows for interrogation of both structural and functional networks at a whole-brain and targeted level, proving to be an excellent method for exploring post-stroke reorganisation and recovery. Given the relative infrequency of childhood AIS and limited studies exploring subsequent functional recovery patterns in this subgroup, the incorporation of children into this study provides particularly valuable insights.

The ability to provide accurate prognostic counselling at stroke diagnosis in both neonates and children continues to be inadequate. As demonstrated, the neuroimaging techniques utilized in this study are powerful tools for interrogating the motor network and have potential to be applied in the clinical setting to improve prognostication. Therapies targeting the affected hemisphere to increase retention of typical contralateral/ipsilesional motor innervations, such as transcranial magnetic stimulation and constraint-based movement therapy, have been applied to both the perinatal and childhood stroke populations with some positive effect on recovery.^9,,37,40^ Our neuroimaging methods may be valuable tools to provide functional baseline measures and subsequent serial assessments of the impact of such therapeutic interventions on structural and functional reorganisation.

Limitations of this study include small participant numbers, heterogenous stroke characteristics including age at stroke/time of imaging and lesion size/ topography, and a lack of a healthy control group. Future research includes investigation of the role other motor network structures play in motor reorganization following perinatal and childhood AIS, with the application of the same neuroimaging techniques utilized in this study to the under-investigated motor-network regions such as the premotor cortex, supplementary motor area and cerebellum.

In conclusion, this study demonstrates that focal brain injury in the perinatal and childhood period provides a valuable opportunity to study developmental neuroplasticity using advanced MR imaging. The variability in motor outcomes following perinatal and childhood stroke reflects important differences in motor network reorganization. The novel imaging techniques utilised in this study, evaluating both structural and functional connectivity, are powerful tools to explore motor network reorganization and recovery in this population. We demonstrated different patterns of cortical activation and WM connectivity, and the utilization of compensatory non-CST WM tracts, following perinatal and childhood stroke, related to motor outcomes. This research has potential to assist prognostication and assess the impact of targeted therapy to improve long-term motor outcomes following stroke in childhood.

## Sources of Funding

Funding for this project was provided by a 2016 grant from the Brain Foundation: *“Brain structural and motor function correlations in childhood arterial ischaemic stroke using multimodal magnetic resonance imaging”*. Dr SRC undertook a paid research fellowship funded by the RCH Foundation to carry out this project.

Drs JYMY and AW acknowledge position funding support from the Royal Children’s Hospital Foundation (RCHF 2022−1402). The funding sources had no role in the study design; in the collection, analysis and interpretation of data; in the writing of the report; and in the decision to submit the article for publication.

## Disclosures

The authors report no disclosures.

## Data Availability

The corresponding author has full access to all the data in the study and takes responsibility for its integrity, analysis and availability.

## Non-standard Abbreviations and Acronyms

AIS: arterial ischemic stroke
ACA: anterior cerebral artery
ASPECTS: Alberta Stroke Programme Early CT Score
BG: basal ganglia
BOLD: blood-oxygen-level-dependent
CC: corpus callosum
CST: corticospinal tract
DWI: diffusion weighted imaging
FEAT: fMRI Expert Analysis Tool
FOD: fibre orientation distributions
GLM: general linear model
ID: identification number
LI: laterality indices
M1: primary motor area
MCA: middle cerebral artery
MCP: middle cerebellar peduncle
MSMT-CSD: multi-tissue Constrained Spherical Deconvolution
PAIS: perinatal arterial ischemic stroke
PCA: posterior cerebral artery
PPERI: presumed perinatal infarction
PSOM: Pediatric Stroke Outcome Measure
ROI: regions of interest
S1: primary somatosensory area
SCP: superior cerebellar peduncle
SCT: striatrocortical tract
SM: sensorimotor
SMA: supplementary motor area
ST_PAR: striato-parietal
ST_PREC: striato-precentral
ST_PREM: striato-premotor
ST_POSTC: striato-postcentral
STR: superior thalamic radiation
T_PAR: thalamo-parietal
T_PREC: thalamo-precentral
T_PREM: thalamo-premotor
T_POSTC: thalamo-postcentral
TCT: thalamocortical tract
Thalm: thalamus
TW-FC: track-weighted functional connectivity
WBT: whole brain tractogram
WM: white matter

## Acknowledgements

This research was conducted within the Department of Neurology, Neurosurgery, The Royal Children’s Hospital, and the Developmental Imaging Group, Murdoch Children’s Research Institute at the Melbourne Children’s MRI Centre, Melbourne, Victoria. It was supported by The Royal Children’s Hospital Foundation, Murdoch Children’s Research Institute, The University of Melbourne, Department of Paediatrics, and the Victorian Government’s Operational Infrastructure Support Program.

We acknowledge the support of Belinda Stojanovski & Adam Rosza (RCH Stroke Research Coordinators), Mr Mike Kean & the RCH imaging team, Dr Sarah Barton, Ms Marilyn Bear, Ms Sue Greaves, Ms Deanne Thompson, Dr Simon Harvey, Dr Adam Scheinberg and Ms Joannah Tozer. We also acknowledge the children and families who generously gave their time to participate in this study.

## Supplementary Material

Figure S1: BOLD-fMRI activation maps for all participants grouped by no hemiparesis/hemiparesis

Figure S2: TW-FC maps for all participants grouped by no hemiparesis/hemiparesis

## Notes

### Competing Interest Statement

The authors have declared no competing interest.

### Author Declarations

The Human Research and Ethics Committee of the Royal Children's Hospital, Melbourne, Australia, gave ethical approval for this work.

